# Surgical site infection and its associated factors in Ethiopia: A systematic review and meta-analysis

**DOI:** 10.1101/2019.12.25.19015792

**Authors:** Wondimeneh Shibabaw Shiferaw, Yared Asmare Aynalem, Tadesse Yirga Akalu, Pammla Margaret Petrucka

**Affiliations:** Lecturer of Nursing, Department of Nursing, College of Health Science, Debre Berhan University; Lecturer of Nursing, Department of Nursing, College of Health Science, Debre Markos University; College of Nursing, University of Saskatchewan

**Keywords:** surgical site infections, wound infection, postoperative, Ethiopia

## Abstract

**Background:** Despite being a preventable complication of surgical procedures, surgical site infections (SSIs) continue to threaten public health with significant impacts on the patients and the health-care human and financial resources. With millions affected globally, there issignificant variation in the primary studies on the prevalence of SSIs in Ethiopia. Therefore, this study aimed to estimate the pooled prevalence of SSI and its associated factors among postoperative patients in Ethiopia.

**Methods:** PubMed, Scopus, Psyinfo, African Journals Online, and Google Scholar were searched for studies that looked at SSI in postoperative patients. A funnel plot and Egger’s regression test were used to determine publication bias. The I^2^ statistic was used to check heterogeneity between the studies. DerSimonian and Laird random-effects model was applied to estimate the pooled effect size, odds ratios (ORs), and 95% confidence interval (CIs) across studies. The subgroup analysis was conducted by region, sample size, and year of publication. Sensitivity analysis was deployed to determine the effect of a single study on the overall estimation. Analysis was done using STATA™ Version 14 software.

**Result:** A total of 24 studies with 13,136 study participants were included in this study. The estimated pooled prevalence of SSI in Ethiopia was 12.3% (95% CI: 10.19, 14.42). Duration of surgery > 1 hour (AOR = 1.78; 95% CI: 1.08 –2.94), diabetes mellitus (AOR = 3.25; 95% CI: 1.51–6.99), American Society of Anaesthesiologists score >1 (AOR = 2.51; 95% CI: 1.07–5.91), previous surgery (AOR = 2.5; 95% CI: 1.77–3.53), clean-contaminated wound (AOR = 2.15; 95% CI: 1.52–3.04), and preoperative hospital stay > 7 day (AOR = 5.76; 95% CI: 1.15–28.86), were significantly associated with SSI.

**Conclusion:** The prevalence of SSI among postoperative patients in Ethiopia remains high with a pooled prevalence of 12.3% in 24 extracted studies. Therefore, situation based interventions and region context-specific preventive strategies should be developed to reduce the prevalence of SSI among postoperative patients.

## Introduction

Surgical site infections (SSIs) are infections that occur at or near surgical incision within 30 days of operation or after 1 year if an implant is placed [1]. SSIs are a major cause of morbidity and mortality worldwide, affecting 5.6% of surgical procedures in developing countries [2]. According to a World Health Organization (WHO) report, the incidence of SSIs ranges from 1.2 to 23.6 per 100 surgical procedures [3]. Worldwide, it has been reported that more than one-third of postoperative deaths are related to SSIs [4]. In addition, SSIs threaten the lives of millions of patients each year and contribute to the spread of antibiotic resistance[5].

The incidence of SSIs is higher in developing countries relative to developed nations [6], reported as the second most common cause of hospital acquired infection (HAI) in Europe and the United States of America (USA) [7]. Approximately 2–5% of surgical patients worldwide have developed SSIs [8]. SSIs are the most frequent type of HAI in low and middle income countries (LMICs) and affect up to one third of patients who have undergone a surgical procedure [9, 10]. In LMICs, the pooled incidence of SSI was 11.8 per 100 surgical procedures [11]. In Africa, SSIs were the leading infections in hospitals and incidence ranged from 2.5%-30.9% [12]. Substantial evidentiary variation on the prevalence of SSI exists across the globe, such as 10.56% in Nnewi [13], 53% in Iran [14], and 16.4% in Uganda [15].

According to recent evidence, the risk factors for SSI are multifactorial and complex. For instance, pre-existing illness [16-18], wound contamination [13, 15, 19], American Society of Anaesthesiologists (ASA) score III or IV [13, 20], non-use of prophylactic antibiotics [21], presence of hypovolemic [22], longer duration of operation [16, 17, 21], longer preoperative hospital stay [16, 21], postoperative hospital stay [16, 19], advanced age [16, 21], alcohol use [19, 23], previous surgery [23], use of drain [17], use of iodine alone in skin preparation [17], smoking [17, 18], absence of wound care [18], and hair removal inside operating room [20] were factors associated with SSIs.

Though SSIs are among the most preventable healthcare-associated infections. However, according to the available global evidence, SSIs impose significant burden to the patient and health care system in terms of prolonged hospital stays [24], spend time in an intensive care unit [7], readmission to hospital [25], long-term disability [26], contribute to spread of antibiotic resistance [3, 11], increase treatment intensity [24], substantial financial burden to health care systems [7, 26], high costs for patients and families [7, 27, 28], deterioration in the quality of life [29], and unnecessary deaths [3, 7, 26]. Effectively controlling SSIs can reduce some of these negative effects [11], as up to one-half of SSIs can generally be prevented through an improved adherence to established basic principles, such as surgical hand preparation, skin antisepsis, adequate antibiotic prophylaxis, less traumatic, less invasive and shorter surgery duration, improved haemostasis and avoidance of hypothermia [7, 23, 30].

Despite improvements in operating room practices, instrument sterilization methods, better surgical technique, and the best efforts of infection prevention strategies, surgical site infections remain a major cause of hospital acquired infections. The need for SSIs prevention policies has been recognized in Ethiopia. Different primary studies in Ethiopia show the magnitude of SSIs as a health issue in the region; however, incidence rates are inconclusive. Therefore, this review and meta-analysis aimed to estimate the pooled national prevalence of SSI and its associated factors in Ethiopia.

## Methods

### Literature search strategy

Initially, Cochrane library, JBI and PROSPERO databases were searched. To confirm whether systematic review and meta-analysis is exist or for the presence of ongoing projects related to the current topic. The literature was searched using PubMed, Scopus, Google scholar, African journals online, and Psyinfo. Relevant articles were identified according to the following terms: “surgical site infections”, “surgical incision infection”, “postoperative”, “wound infection”, “predictor”, “associated factors”, “Ethiopia”. The key terms were used in combination using Boolean operators like “OR” or “AND”. The searches were restricted to full texts, free articles, human studies, and English language publications. This search involved articles published from January 1^st^, 2000 to November 11^th^, 2019. Grey literatures, such as surveillance reports, academic dissertations, conference abstracts, were examined. In addition the reference lists of included articles were hand-searched to identify any potential additional relevant articles.

### Eligibility criteria

Studies were included in the meta-analysis if they adhered to the following guidelines: (1) all observational study designs (cross-sectional, case-control, and cohort studies) needed to report the prevalence of SSI; (2 published from 2000 to 2019; (3) published in English language; (4) abstract and full text were available for this review; and (5) conducted in Ethiopia. Studies were excluded if they: (1) possessed a poor quality score as per the stated criteria; (2) failed to determine the desired outcome (SSI); (3) were not fully accessible; and (4) included only in cesarean section patients.

### Outcome of interest

The main outcome of interest was the prevalence of SSIs reported in the original paper both as percentage or as the number of SSIs cases(n) / total number of patients who undergo surgery (N). These two parameters were necessary to calculate the pooled prevalence of SSIs in the meta-analysis. Therefore, the prevalence was calculated by dividing the number of individuals who have SSIs to the total number of patients who undergo surgery (sample size) multiplied by 100.

### Data extraction

Two authors independently extracted all necessary data from each study using a standardized data extraction format. If discrepancies between data extractors were observed, a third author was involved. For each included study, the following data were extracted: primary author, publication year, region, study design, sample size, prevalence of SSIs, and associated factors (wound type, preoperative hospital stay, duration of operation, history surgery, ASA score, diabetes mellitus, smoking, preoperative blood transfusion). We also contacted corresponding authors (by e-mail) of articles that did not provide details of their study’s background and asked them for the relevant information, such as study time, region, or hospital.

### Quality assessment

Two independent investigators assessed the methodological quality of all of the potential studies to be included in our analysis. Any disagreements between the authors were resolved through discussion or, if consensus could not be reached, consultation with a third, independent author was undertaken. The quality of each included study was assessed using the Joanna Briggs Institute (JBI) quality appraisal checklist [31]. This scale has several key criteria to appraise cohort studies including: [1] similarity of groups, [2] similarity of exposure measurement, [3] validity and reliability of measurement, [4] identification of confounder, [5] strategies to deal with confounder, [6] appropriateness of groups/participants at the start of the study, [7] validity and reliability of outcome measured, [8] sufficiency of follow-up time, [9] completeness of follow-up or descriptions of reason to loss to follow-up, [10] strategies to address incomplete follow-up, and [11] appropriateness of statistical analysis. The items used to appraise cross-sectional studies were: [1] inclusion criteria, [2] description of study subject and setting, [3] valid and reliable measurement of exposure, [4] objective and standard criteria used, [5] identification of confounder, [6] strategies to handle confounder, [7] outcome measurement, and [8] appropriate statistical analysis. Studies were considered low risk when scored at 50% and above on the quality assessment indicators.

### Statistical analysis

To obtain the pooled prevalence of surgical site infection, a meta-analysis using random-effects DerSimonian and Laird model was performed due to anticipated heterogeneity [32]. Cochran’s Q chi-square statistics and I^2^ statistical test was conducted to assess the random variations between primary studies [33]. In this study, heterogeneity was interpreted as an I^2^ value of 0% = no heterogeneity, 25% = low, 50% = moderate, and 75% = high [34]. In case of high heterogeneity, subgroup analysis and sensitivity analyses were run to identify possible moderators of this heterogeneity. Potential publication bias was assessed by visually inspecting funnel plots and objectively using the Egger bias test [35]. To account for any publication bias, we used the trim-and-fill method, based on the assumption that the effect sizes of all the studies are normally distributed around the center of a funnel plot. The meta-analysis was performed using the STATA™ Version 14 software [36]. Finally, for all analyses, P < 0.05 was considered statistically significant.

### Presentation and reporting of results

The results of this review were reported based on the Preferred Reporting Items for Systematic Review and Meta-Analysis statement (PRISMA) guideline [37]. (Supplementary file-PRISMA checklist). The entire process of study screening, selection, and inclusion were shown with the support of a flow diagram. Additionally, tables and narrative summaries were used to report the risk of bias for every eligible study.

## Results Search results

In the first step of our search, 293 studies were retrieved. About 289 studies were found from five international databases and the remaining 4 were through a manual search. The databases included PubMed (45), Scopus (31), Google Scholar (152), Psyinfo (29) and African Journals Online (59). Of these, 128 duplicate records were identified and removed. From the remaining 168 articles, 113 articles were excluded after reading the titles and abstracts based on the pre-defined eligibility criteria. Finally, 55 full-text articles were assessed for their eligibility. Based on the pre-defined criteria and quality assessment, 24 articles were included for the final analysis. The detailed steps of the screening process are shown in a PRISMA flow chart of the study selection (Fig. 1).

**Figure 1.**
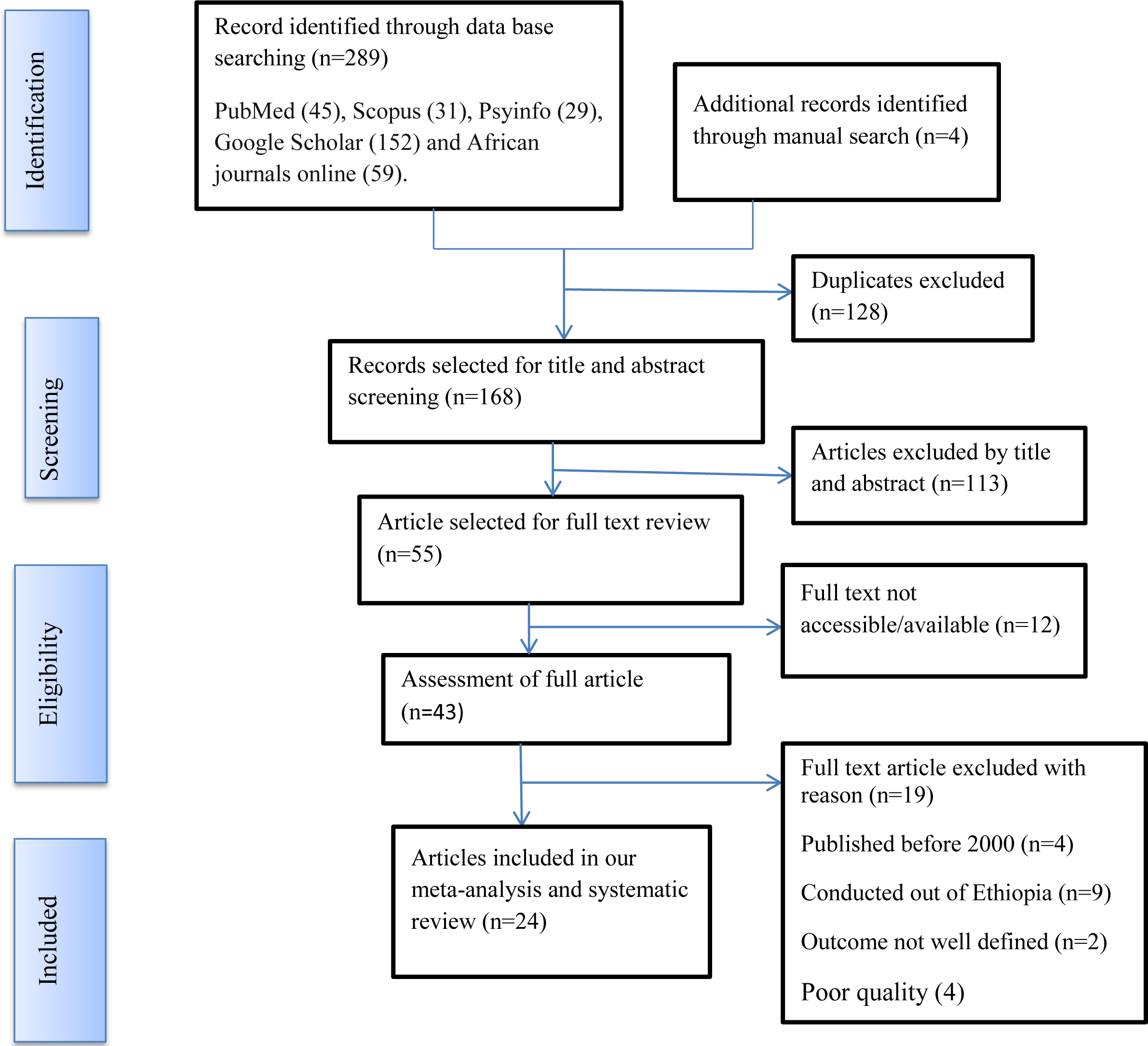
PIRSMA Flowchart diagram of the study selection

### Baseline characteristics of included studies

In the current meta-analysis, a total of 24 studies with 13,136 study participants were included to estimate the pooled prevalence of SSIs among post-operative patients. With respect to design, two thirds (62.5%) of the studies included were cross-sectional. The studies varied substantially in sample size, ranging from 65 to 1754. The highest prevalence (36%) of SSIs was reported in a study conducted in Oromia [38]. Whereas the lowest prevalence (3.5%) was reported in a study conducted in Amhara [39]. Overall information regarding the prevalence of SSIs in post-operative patients was obtained from various regions in Ethiopia. Seven of the studies involved participants from Addis Ababa [23, 40-45], eight from Amhara [16, 18, 39, 46-50, four from Oromia [38, 51-53], three from Tigray [19, 54, 55], and two from SNNPR [21, 56]. Regarding the sampling technique, 12 studies [16, 21, 38, 39, 42, 47, 49-52, 54, 56] used the consecutive sampling technique to select study participants. However, two studies [44, 55] did not report their sampling methods. The quality score of each primary study, based on the JBI quality appraisal criteria, showed no considerable risk; hence, all the studies were considered in this review (Table 1).

**Table 1.** Baseline characteristics of the included studies

| Author | Publication year | Region | Study design | Sample size | Prevalence (%) | Data collection method | Sampling technique | Study period | Quality Score |
| --- | --- | --- | --- | --- | --- | --- | --- | --- | --- |
| Afenigus.A.et al [18] | 2019 | Amhara | Cross-sectional | 165 | 25.5 | Interview and clinical sign | Systematic | March to May, 2017 | Low risk |
| Alamrew K. [40] | 2018 | Addis Ababa | Cross-sectional | 413 | 11.1 | Chart review | Simple random | June 10 to September 10, 2016 | Low risk |
| Ali S.et al [51] | 2018 | Oromia | Cohort | 1069 | 13.85 | Laboratory and interview | Consecutive | May to September, 2016 | Low risk |
| Amare B [39] | 2011 | Amhara | Cross-sectional | 1627 | 3.5 | Clinical sign | Consecutive | January 2010 to June 2010 | Low risk |
| Amenu D. et al [52] | 2011 | Oromia | Cross-sectional | 770 | 11.4 | Interview, logbook | Consecutive | April 1, 2009 to March 31 2010 | Low risk |
| Argaw NA. et al [41] | 2017 | Addis Ababa | Cross-sectional | 200 | 16 | Chart review | All included | NR | Low risk |
| Billoro BB. et al [56] | 2019 | SNNPR | cohort | 280 | 16.5 | Interview, chart review, observation, laboratory | Consecutive | January 1 to September 1, 2017 | Low risk |
| Endalafer N. [42] | 2011 | Addis Ababa | Cross-sectional | 215 | 17.67 | Laboratory | Consecutive | June 2007 to April 2008 | Low risk |
| Fisha K. et al [46] | 2019 | Amhara | Cohort | 642 | 9.9 | Chart review | Simple random | March 15 to April 15, 2018 | Low risk |
| Forrester JA. et al [38] | 2018 | Oromia | Cohort | 65 | 36 | Clinical sign | Consecutive | September 9 to 20, 2016 | Low risk |
| Gebremeskel S. [43] | 2018 | Addis Ababa | Cross-sectional | 410 | 4.8 | Chart review, interview and laboratory | Systematic | March 1 to August 30, 2017 | Low risk |
| Gedefaw.G. et al [47] | 2018 | Amhara | Cross-sectional | 447 | 9.6 | Chart review | Consecutive | March 1 to April 30, 2018 | Low risk |
| Gelaw A. et al [48] | 2014 | Amhara | Cross-sectional | 510 | 8.2 | Laboratory | All included | November 2010 to February 2011 | Low risk |
| Halawi.E. et al [23] | 2018 | Addis Ababa | Cohort | 158 | 20.6 | Clinical sign, chart review, laboratory | Convenient | April 1 to April 30, 2017 | Low risk |
| Laloto TL. et al [21] | 2017 | SNNPR | Cohort | 127 | 19.1 | Chart review, clinical sign, interview | Consecutive | March 2 to May 2, 2015 | Low risk |
| Mamo T.et al [53] | 2017 | Oromia | Cohort | 390 | 9.4 | Laboratory, chart review, interview | Purposive | April 23 to September 5, 2015 | Low risk |
| Melku S.et al [49] | 2012 | Amhara | Cross-sectional | 1383 | 8 | Interview, chart review, laboratory | Consecutive | April to August 2009 | Low risk |
| Mengesha RE.et al [54] | 2014 | Tigray | Cross-sectional | 610 | 20.9 | Clinical sign and chart review | Consecutive | January to June 2012 | Low risk |
| Mulu W. et al [16] | 2013 | Amhara | Cross-sectional | 294 | 10.2 | Clinical sign, culture, and chart review | Consecutive | October 2010 to January 2011 | Low risk |
| Taye M. [44] | 2005 | Addis Ababa | Cohort | 1754 | 14.8 | Clinical sign and laboratory | NR | January 1, 1999 to Dec 31, 1999 | Low risk |
| Tekie K. [45] | 2008 | Addis Ababa | Cohort | 173 | 17.9 | Clinical sign and laboratory | All included | April to July 2006 | Low risk |
| Tesfahunegn Z. et al [55] | 2009 | Tigray | Cross-sectional | 246 | 7.3 | Chart review and clinical sign | NR | November 2005 to April 2006 | Low risk |
| Weldu MG. et al [19] | 2018 | Tigray | Cross-sectional | 280 | 11.1 | Interviewer, clinical sign | Simple random | February 2 to March 31, 2016 | Low risk |
| Yallew WW. Et al [50] | 2016 | Amhara | Cross-sectional | 908 | 7.6 | Clinical sign, laboratory, chart review | Consecutive | March to July 2015 | Low risk |

**NR:** non-reported

### Prevalence of surgical site infection

The current meta-analysis using the random effects model showed that the estimated overall prevalence of SSI in post-operative patients was 12.3% (95% CI: 10.19,14.42) with a significant level of heterogeneity (I^2^ = 93.8%; p < 0.001) (Figure 2).

**Figure 2.**
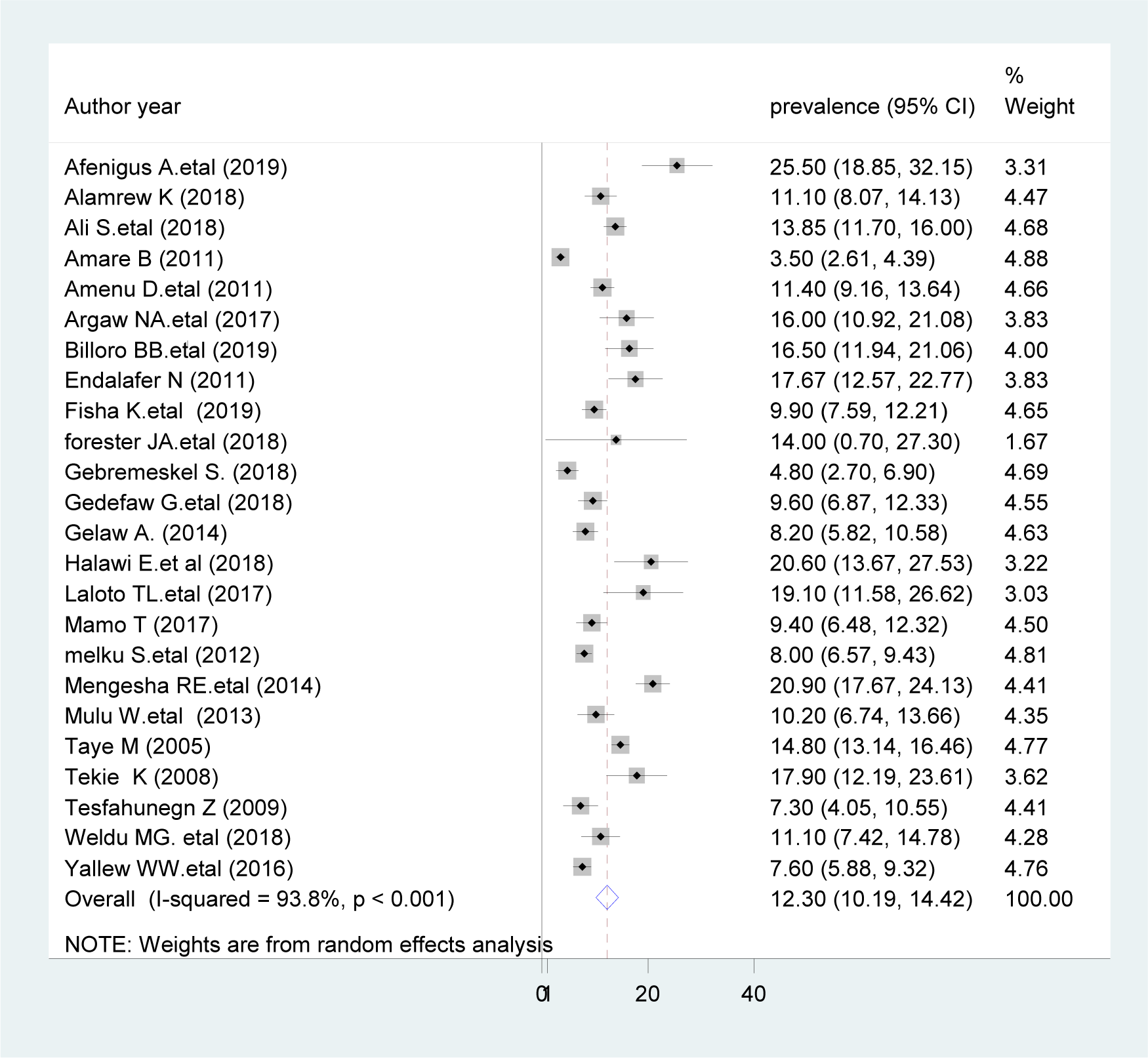
Forest plot of the prevalence of surgical site infections in post-operative patients

### Subgroup analysis

The presence of significant heterogeneity among the primary studies requires the need to conduct subgroup analysis. In order to identify the sources of heterogeneity, we deployed sub-group analysis using publication year, sample size, region, study design, and sampling technique to determine the pooled prevalence of SSIs (Table 2). The prevalence of SSIs was found to be 17.19% in SSNPR, 13.08% in studies published since 2010, 14.18% in studies with cohort design, 15.54% a sample size less than or equal to 300, and 13.38% in studies with probability sampling technique of our included primary studies.

**Table 2.**
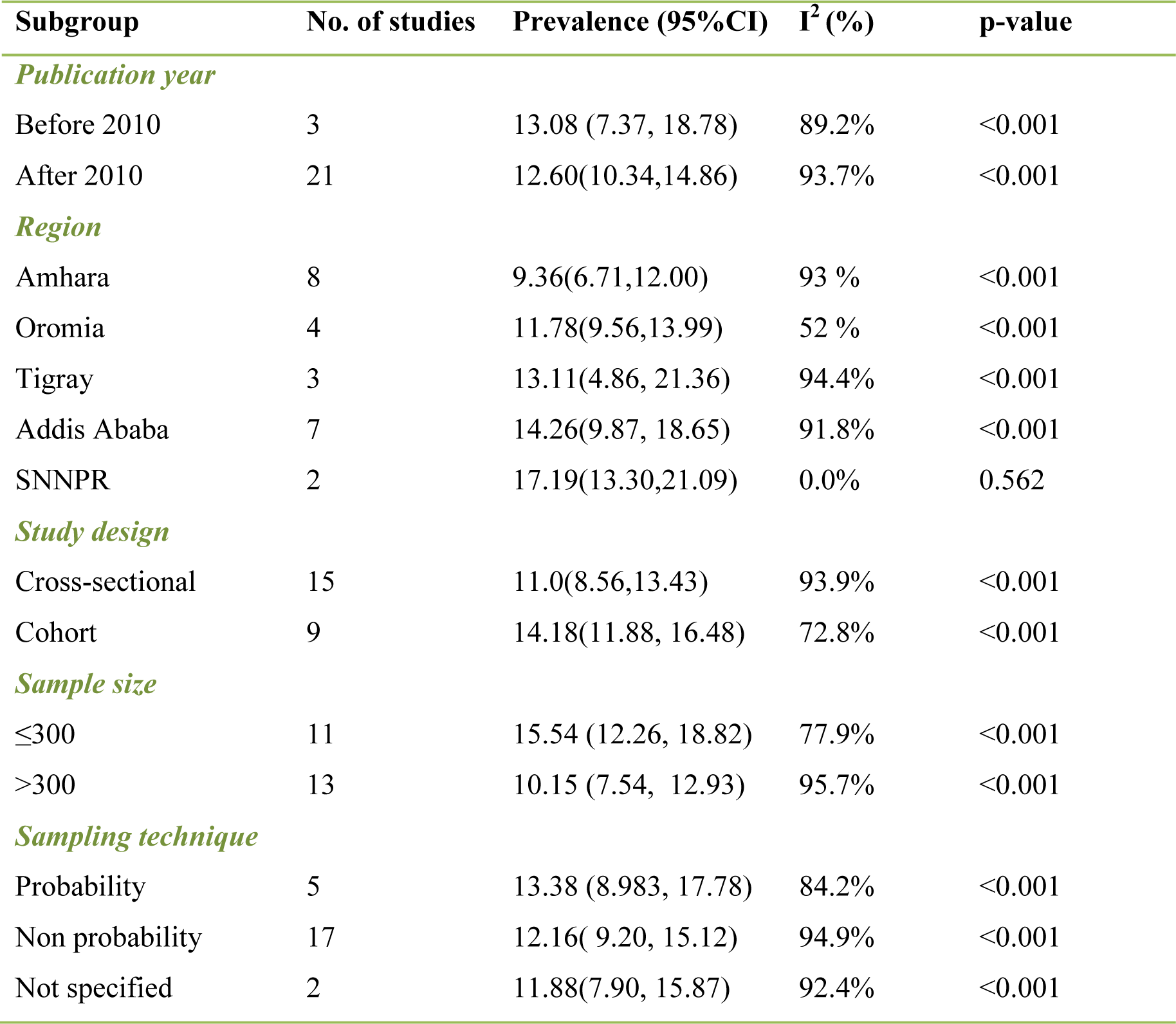
The results of subgroup analysis by characteristics of the studies

### Publication bias

We found that there was publication bias among the included studies, as depicted by the asymmetrical distribution of our funnel plot (Fig. 4). Likewise, the result of Egger’s test was statistically significant for the presence of publication bias (P = 0.001). In addition, to reduce and adjust publication bias, trim and fill analysis was also performed (Figure 5). Trim and fill analysis is a nonparametric method for estimating the number of missing studies that might exist.

**Figure 3.**
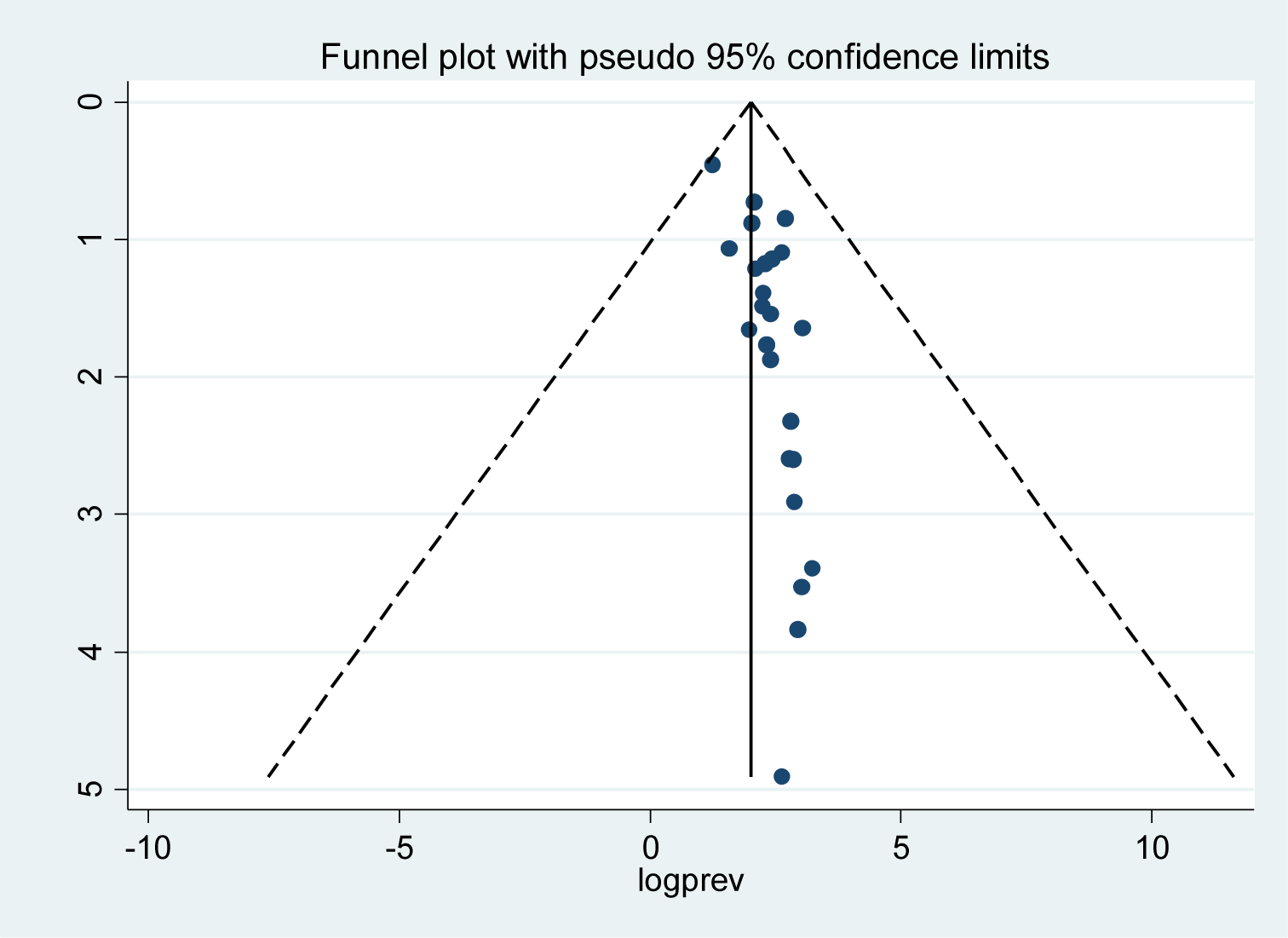
Funnel plot to test publication bias of the 24 studies.

**Figure 4.**
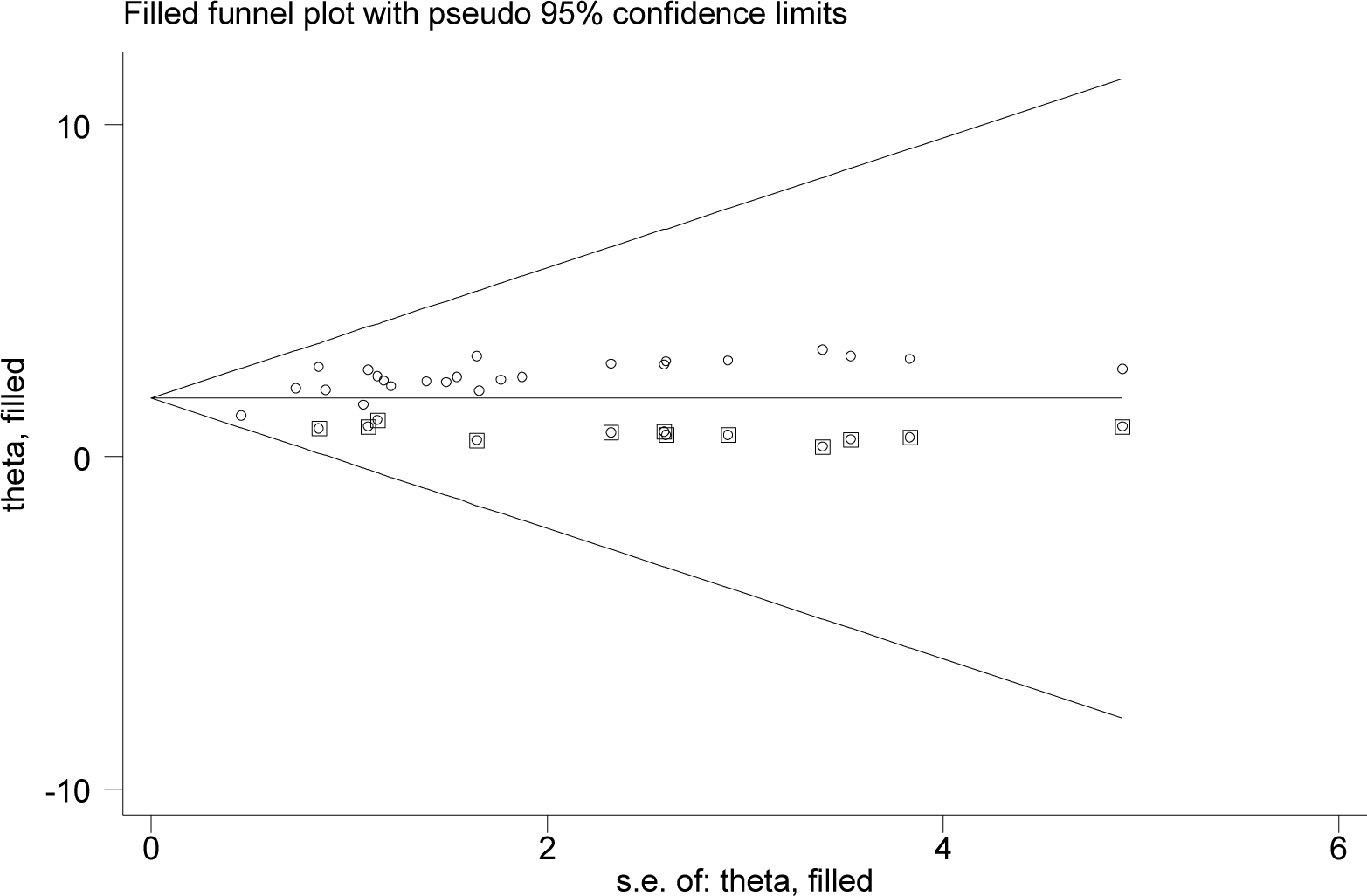
Result of trim and filled analysis for adjusting publication bias of the 24 studies.

**Figure 5.**
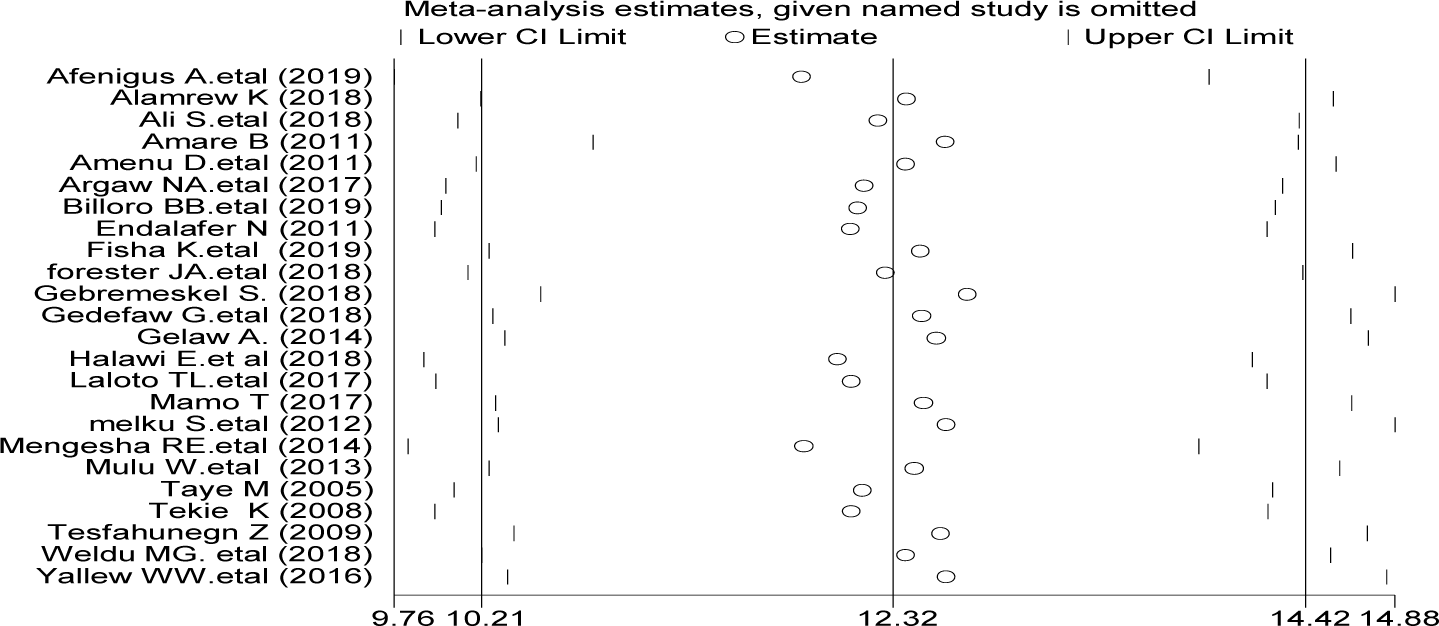
Result of sensitivity analysis of the 24 studies

### Sensitivity analysis

To evaluate the effect of an individual study on the pooled effect size, sensitivity analysis was conducted. Sensitivity analyses using the random effects model revealed that no single study influenced the overall prevalence of SSIs in post-operative patients (Fig. 6)

**Figure 6.**
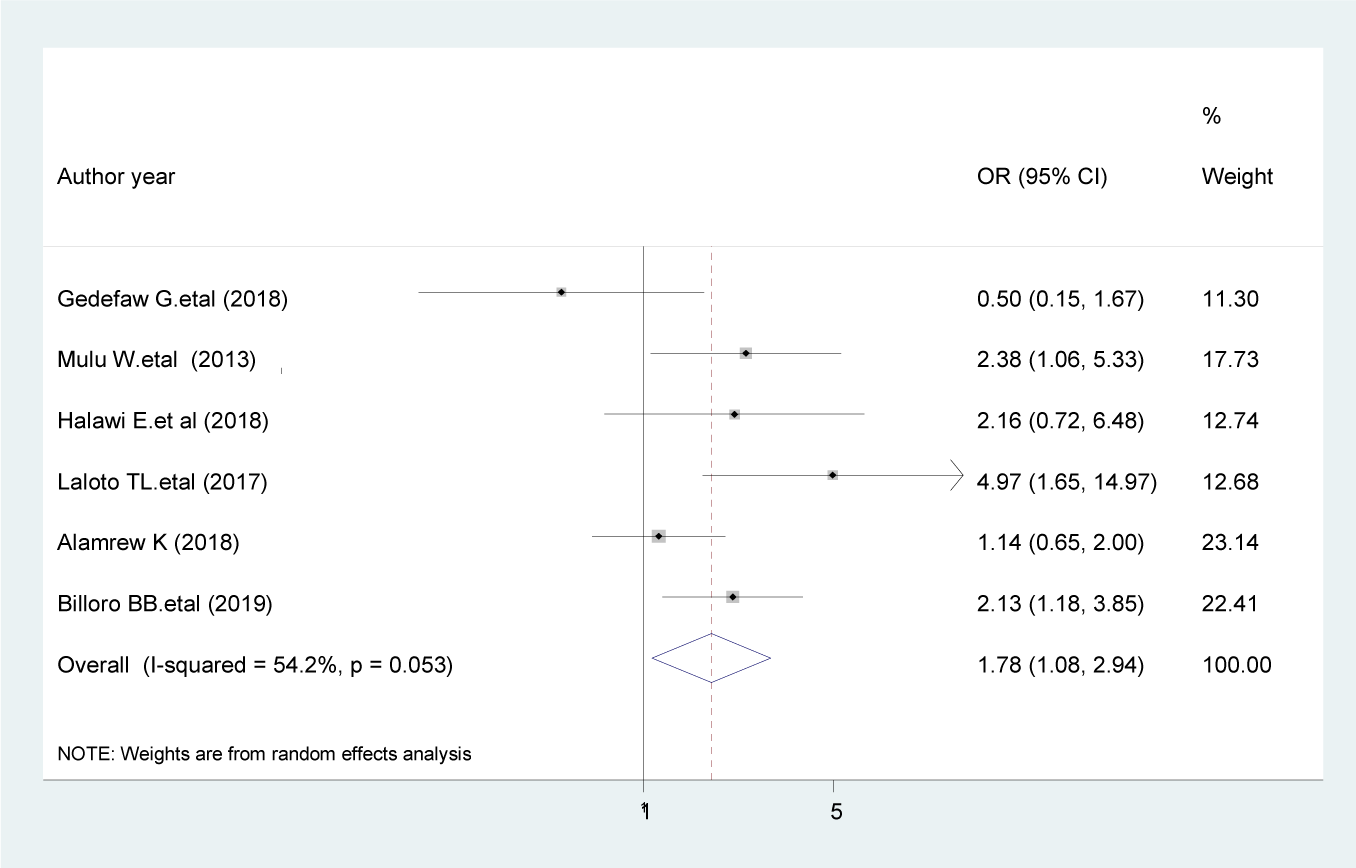
The pooled effect of duration of operation >1 hour on surgical site infection

### Associated factors

Based on this meta-analysis, SSIs in the Ethiopian context were associated with duration of operation, smoking, preoperative blood transfusion, diabetes mellitus, ASA score, previous surgery, wound type, and preoperative hospital stay.

### Duration of operation

Based on the pooled effects of six studies [16, 21, 23, 40, 47, 56], duration of operation greater than one hour were nearly two times more likely to develop SSI as compared with patients whose operation was completed within one hour (AOR, 1.78, 95% CI 1.08, 2.94, I ^2^ =54.2%) (Figure 6). The evidence from Egger’s regression test shows that there was no publication bias (P = 0.803).

### Preoperative blood transfusion

According to our current meta-analysis, for those clients who had preoperative blood transfusion were 2.51 times more likely to develop SSIs as compared with patients with no preoperative blood transfusion (AOR, 2.51, 95% CI 0.67, 9.45, I^2^ =62 %), although this association was not statistically significant (Figure 7). The result of the Egger’s regression test showed no evidence of publication bias (P = 0.838).

**Figure 7.**
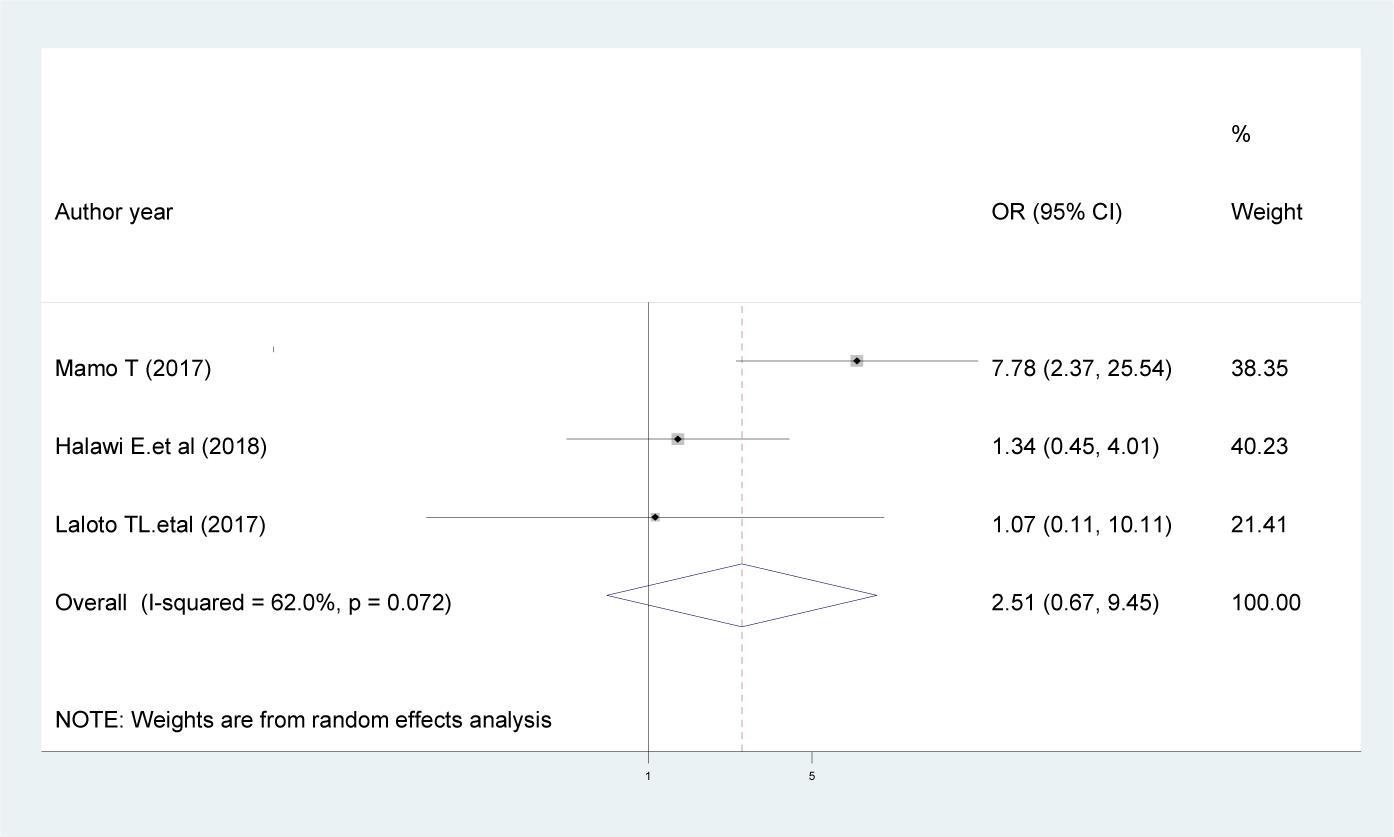
Forest plot showing the association of preoperative blood transfusion and surgical site infection

### Cigarette smoking

Those patients who smoke cigarettes were 95% more likely to develop SSI compared with patients who do not smoke cigarettes [OR = 1.95; 95% CI: 0.27, 13.18, I^2^ =0.0%], though not statistically significant (Fig. 8). The evidence from Egger’s regression test showed that there was no publication bias (P = 0.107).

**Figure 8.**
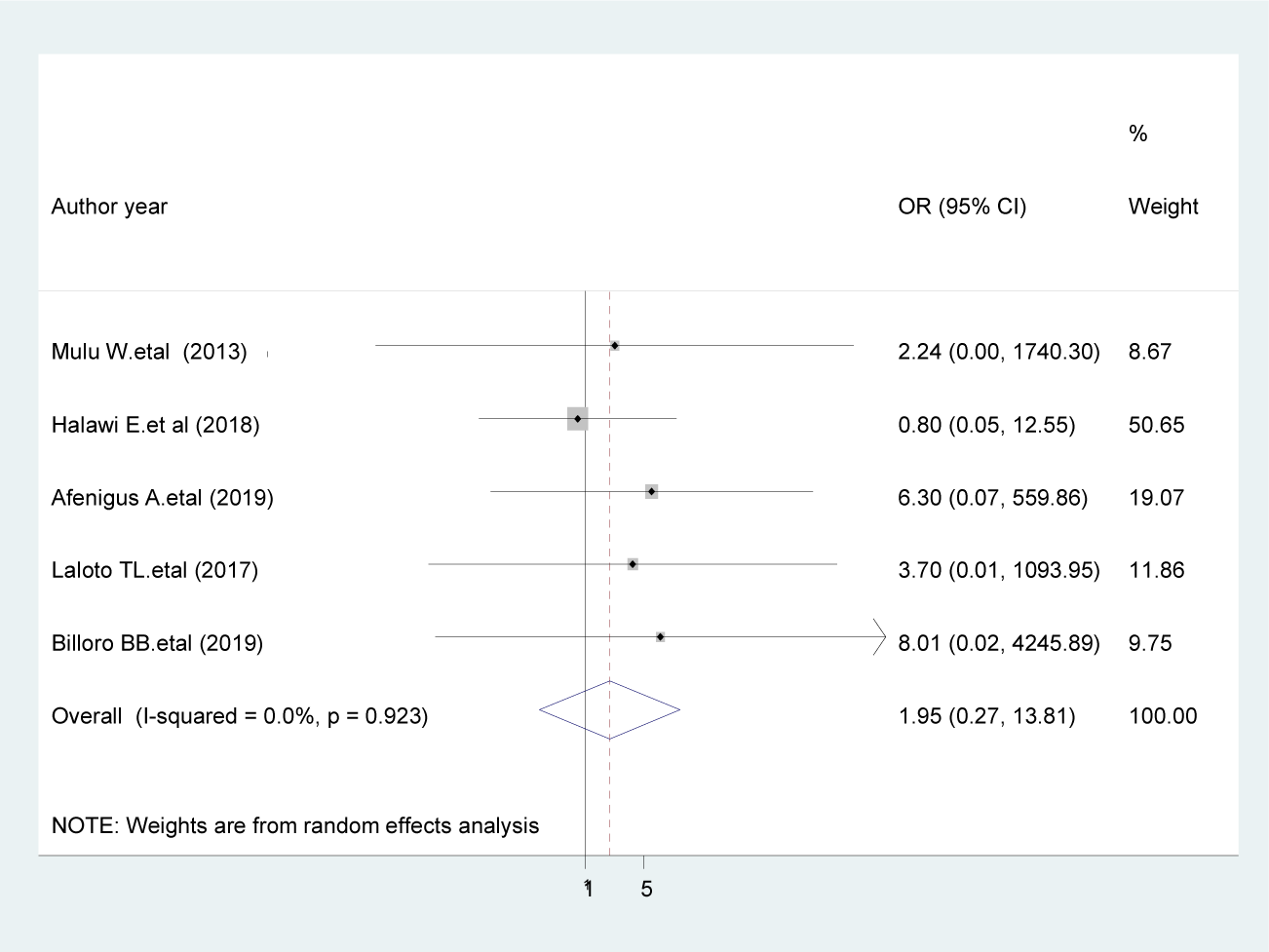
The pooled effect of smoking on surgical site infection

### Diabetes mellitus

The pooled effects of five studies [16, 18, 21, 47, 53] indicated that those patients who had diabetes mellitus at the time of surgery were 3.25 time more likely to develop SSIs than non-diabetic patients [AOR = 3.25; 95% CI: 1.51, 6.99, I^2^ =21.6%] (Figure 9). The evidence from Egger’s test shows no significant proof of publication bias (P = 0.429).

**Figure 9.**
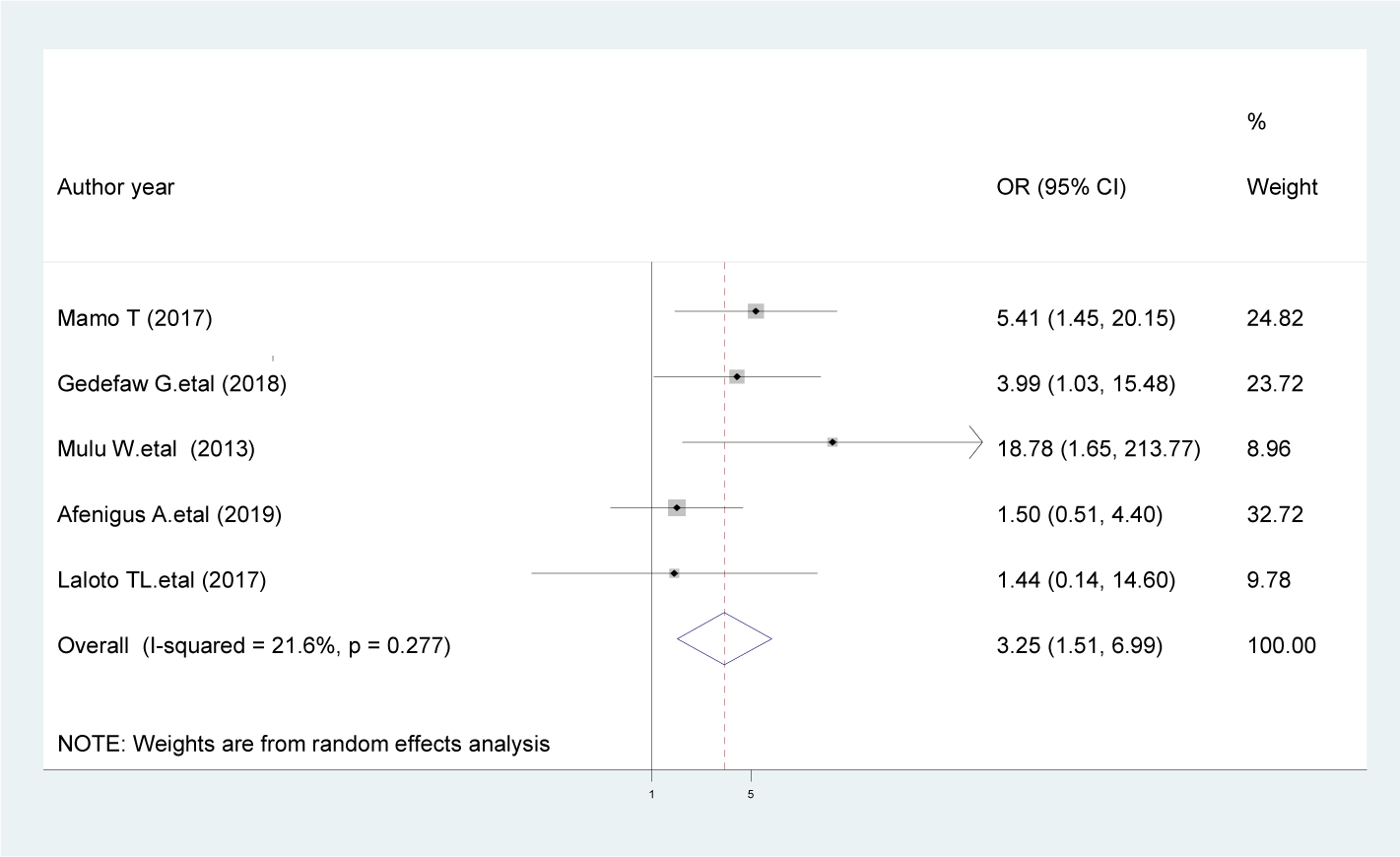
The pooled effect of diabetes mellitus on surgical site infection

### ASA score

The present review revealed that patients with ASA score >1 were 2.51 times more likely to develop SSIs compared with ASA score ≤ 1 [AOR = 2.51; 95% CI: 1.07, 5.91, I^2^ = 0 %] (Figure 10). The evidence from Egger’s test shows no significant proof of publication bias (P = 0.427).

**Figure 10.**
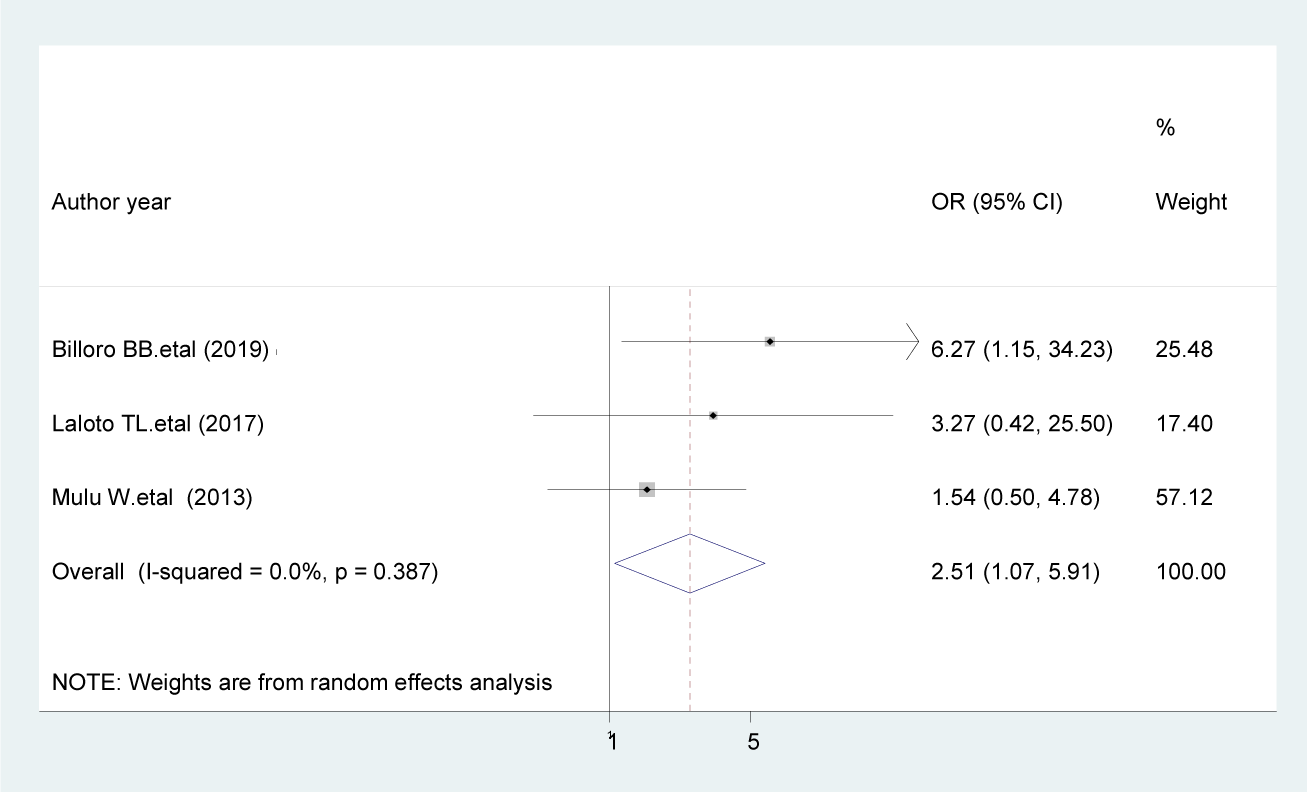
The pooled effect of ASA score on surgical site infection

### Previous surgery

Patients with a history of previous surgery were 2.5 times more likely to develop SSIs compared with patients having no history of previous surgery [AOR = 2.5; 95% CI: 1.77, 3.53, I^2^ = 0 %] (Fig. 11). The result of the Egger’s regression test showed no evidence of publication bias (P = 0.071).

**Figure 11.**
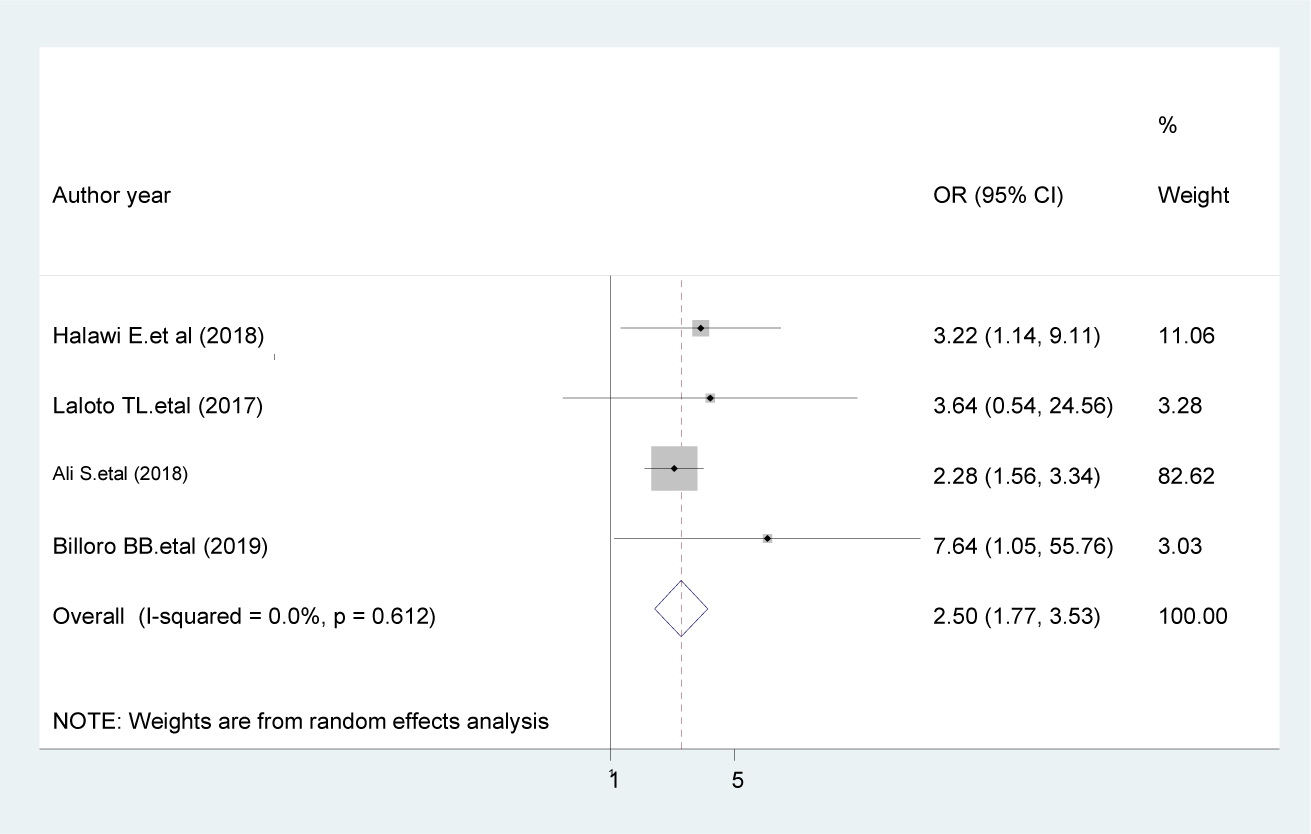
The pooled effect of previous surgery on surgical site infection

### Wound type

The pooled effects of six studies [21, 23, 40, 41, 51, 56] showed that those patients who have clean-contaminated wound were 2.15 time more likely to develop SSIs than those who have clean wound [AOR = 2.15; 95% CI: 1.52, 3.04, I^2^ =0 %] (Fig. 12). The evidence from Egger’s regression test showed that there was no publication bias (P = 0.15).

**Figure 12.**
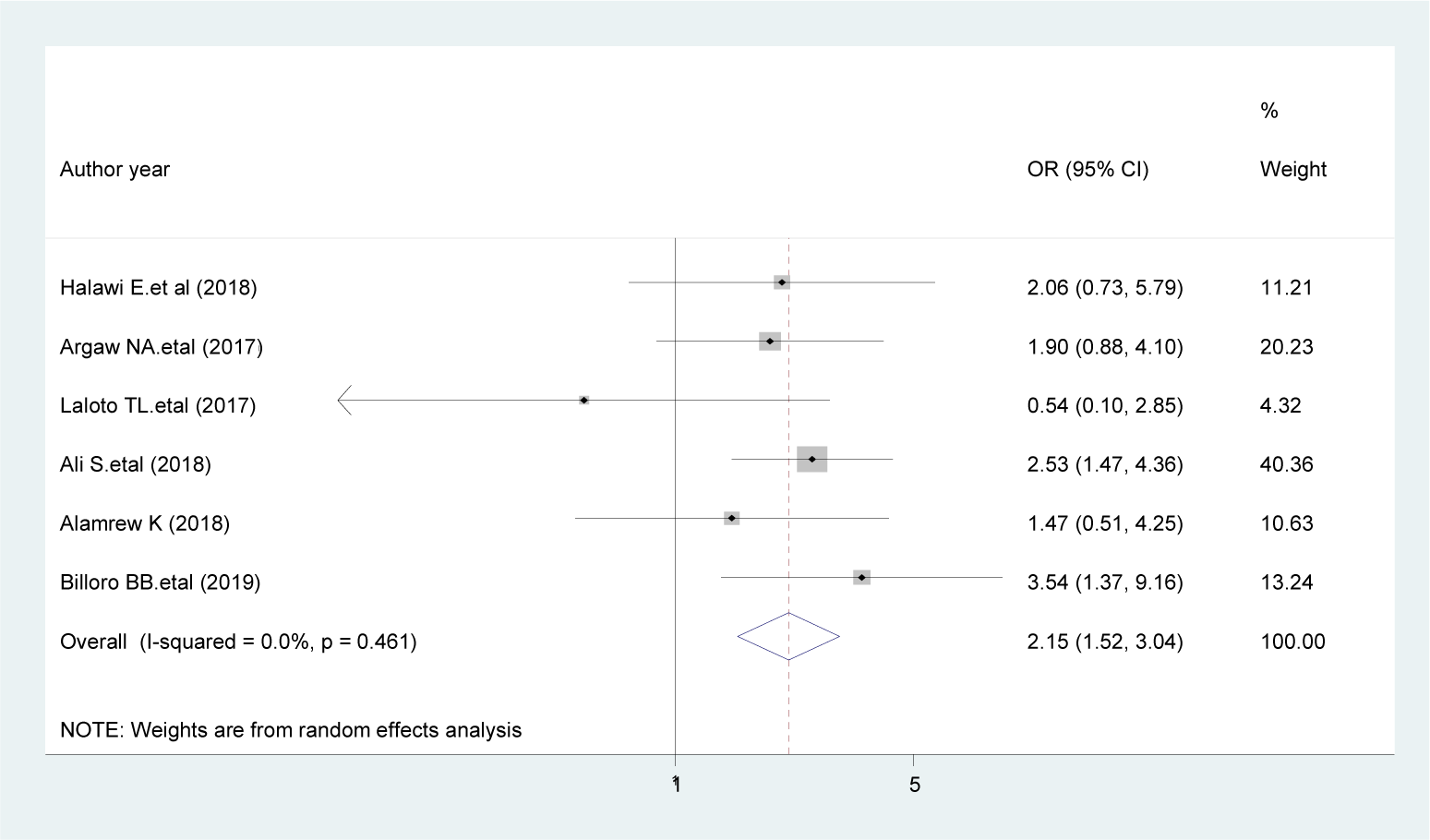
The pooled effect of wound type on surgical site infection

### Preoperative hospital stay

The current meta-analysis showed that patients with preoperative hospital stays greater than 7 days were 5.76 times more likely to develop SSIs compared with patients whose preoperative hospital stays were less than or equal to 7 days [AOR = 5.76; 95% CI: 1.15, 28.86, I^2^ =84.6 %] (Fig. 13). The evidence from Egger’s regression test shows that there was no publication bias (P = 0.545).

**Figure 13.**
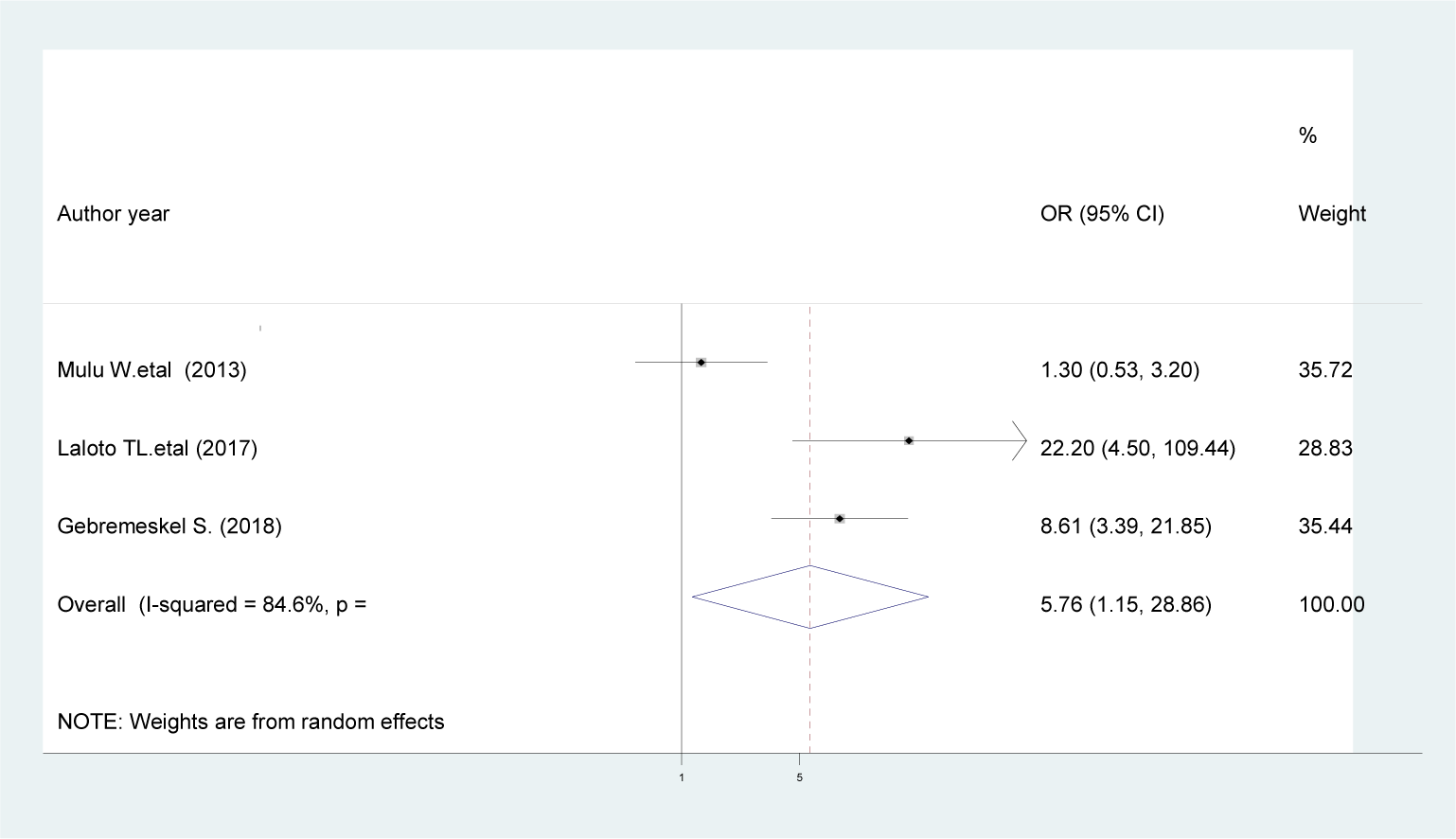
The pooled effect of preoperative hospital stays on surgical site infection

## Discussion

In this meta-analysis the national pooled prevalence of SSIs in Ethiopia was estimated to be 12.3% (10.19–14.42%). This finding indicates that SSIs are highly prevalent in hospital patients and reflects inadequate implementation of infection prevention in Ethiopia. Hence, a multifactorial approach is required to manage SSIs, with emphasis being placed on adequate antibiotic prophylaxis, aseptic wound care, and treatment adherence.

Our estimated prevalence of SSIs among post-operative patients in Ethiopia is in line with systematic review and meta-analysis studies done in Nigeria 14.5% [57] and 14.8% in Sub-Saharan Africa [58]. However, this result was substantially higher than studies conducted in China was 4.5% [59] and 5.4% in Algeria [60]. This variation could be justified by lack of adequate postoperative care, inadequate infection control, and insufficient trained professionals in Ethiopia.

Subgroup analysis in this study showed that the pooled prevalence of SSIs among post-operative patients in SNNPR was 17.19% (95% CI: 13.3, 21.09), which was the highest amongst the Ethiopian regions examined, followed by Addis Ababa (14.26%; 95% CI: 9.87, 18.65), whereas the lowest rate was reported in the Amhara Region (9.36%; 95%CI;6.71, 12.0) of the country. This variation might be due the availability of healthcare resources in the regions and the diagnostic methods used. Based on the pooled analysis of adjusted odd ratio of studies, duration of operation > 1 hour, diabetes mellitus, ASA score >1, previous surgery, clean contaminated wound, and preoperative hospital stay > 7 days were associated with SSIs.

The present study revealed that the duration of operation > 1 hour were nearly two times more at risk to acquire SSIs. This is supported by other studies conducted in Tanzania [17], Nigeria [57], and Spain [61]. Longer duration of surgery would, therefore, increase the risk of surgical wound contamination due to the increased microbial exposure in the operation field [62] and it also increases the extent of tissue trauma due to an extensive surgical procedure and increased blood loss which contributes to tissue hypoxia. Similarly, we found that those who had diabetes mellitus were significantly associated with SSIs. This finding supported by other studies conducted in Yemen [63], Nigeria [57], India [64], and Tanzania [17]. Previous studies have shown that patients with pre-existing illnesses, such as diabetes mellitus are at high risk of developing SSI due to their low immunity and that may slow the healing process [65]. We found that patients who have ASA score >1 were 2.5 times more likely to develop SSIs, which was supported by studies from Canada [66], Uganda [15], Nepal [67], and Cameroon [20].

Those patients with a history of previous surgery had a significant association with a surgical site infection. This finding supported by other studies conducted in India [64]. The odds of clean contaminated wound were nearly two times to have SSI, which is similar to studies from Cameroon [20], China [59], Nepal [67], and Uganda [15]. The possible explanation might be previous studies have shown that ceftriaxone is highly sensitive to suspected organisms in clean wound [56], and surgeries with an increased microbial load in the operative field are associated with higher risk of SSI.

In the present review, preoperative hospital stay for more than 7 days increased the risk of SSIs by 5.76 times compared with preoperative hospital stay less than 7 days. In agreement with this finding were studies conducted in Tanzania [17] and India [64]. The possible explanation might be patients with long stay in hospital before surgery exposes to contamination or colonization by pathogens which will contribute to the occurrence of SSIs [68], suggesting that shortening the preoperative hospital stay reduces the incidence rate of SSIs. Although preoperative blood transfusions and smoking have been established as risk factors for SSIs in other studies [17, 64], these findings were not supported by our meta-analysis results. Further studies are, therefore, required to validate these observations among Ethiopian patients.

This study has clinical implications in that the high prevalence of SSIs among postoperative patients should guide healthcare professionals to minimize the risk of surgical site infection by providing guidance to the patient who undergone to surgery, give information about possible risk factors during routine patient care, and provide knowledge about wound care. The Nurses’ role is considered crucial in optimizing the healing outcomes through aseptic wound care; provision of explicit patient instructions on how to care for their wound, education and counseling and understanding of the patient’s needs. In addition, identifying associated risk factors may help health care professionals treat SSIs patients during their clinical care.

The meta-analysis conducted in this study has limitations that should be considered in future research. First, it is difficult to determine if the results from various region arerepresentative of the entire country, as no data were found for all region of Ethiopia. Second, we did not identify all of the potential predictors of SSIs among postoperative patients. Therefore, further study to identify associated factors for the development of SSI among postoperative patients may be necessary.

## Conclusion and recommendation

This study revealed that the prevalence of SSIs remains high among postoperative patients in Ethiopia based on the 24 research-based papers included in this study. That said, the prevalence of SSIs differed by region. Therefore, situation-based interventions and region context-specific preventive strategies should be developed to reduce the prevalence of SSI. A more comprehensive consideration of the existing evidence will potentially inform effective strategies for preventing SSI within the Ethiopian context. In addition, this meta-analysis may help policy-makers and program managers to design interventions on preventing the occurrence of SSIs.

## Data Availability

which is declared in the main document

## Abbreviations

ASA: American Society of Anaesthesiologists
CI: Confidence Interval
AOR: Adjusted Odds Ratio
PRISMA: Preferred Reporting Items for Systematic Reviews and Meta-Analyses
SNNPR: Southern Nations
Nationalities
and Peoples Region
SSI: Surgical Site Infection.

## Declarations

### Ethics approval and consent to participate

Not applicable.

### Consent for publication

Not applicable.

### Availability of data and materials

The data analysed during the current meta-analysis is available from the corresponding author on reasonable request.

### Authors’ contributions

WSS and TYA developed the protocol and were involved in the design, selection of study, data extraction, statistical analysis, and developing the initial drafts of the manuscript. YAA, TYA and PMP were involved in data extraction, quality assessment, statistical analysis and revising. WSS, YAA, and PMP prepared and edited the final draft of the manuscript. All authors read and approved the final draft of the manuscript.

